# Enhanced Detection Rate of AI for Lung Cancer Detection on GP-Referred Chest X-rays: A Real-World Retrospective Evaluation

**DOI:** 10.64898/2025.12.05.25341684

**Authors:** Anubhav Datta, Pawel Rozwadowski, Philip Broadhurst, Matthew Evison, Anna Sharman, Rhidian Bramley

**Affiliations:** Clinical Radiology, The Christie NHS Foundation Trust; Division of Cancer Sciences, The University of Manchester; Clinical Radiology, Manchester University NHS Foundation Trust; Respiratory Medicine, Manchester University NHS Foundation Trust

**Author notes:** joint first authors (Anubhav Datta, Pawel Rozwadowski). Correspondence, Postal address : Dr Rhidian Bramley, Clinical Radiology, The Christie NHS Foundation Trust, 550 Wilmslow Road, Manchester M20 4BX. United Kingdom.

**Keywords:** Chest radiography, Lung cancer, Artificial intelligence, Computer-aided detection, Diagnostic errors, Missed diagnosis

## Abstract

**Objectives:** To assess whether an artificial intelligence (AI) chest radiograph (CXR) tool could enhance lung cancer detection on primary care–referred CXRs in the UK, and to estimate the magnitude of any improvement.

**Methods:** From ∼280,000 primary care–referred CXRs, we identified 1,600 linked to a lung cancer diagnosis (ICD-10 C34) within six months. Missed lung cancers were defined by review of the CXR report and comparison of diagnostic CT and positron emission tomography (PET) imaging with the index CXR by three specialist radiology clinicians. CXRs with a retrospectively visible but initially missed cancer were re-analysed using a commercially available AI tool. The primary outcome was the enhanced detection rate (EDR), defined as the proportion of confirmed cancers missed on CXR but correctly identified by AI.

**Results:** Of 1,600 CXRs, 105 (6.6%) contained a retrospectively visible cancer that had been missed at first report. AI flagged abnormalities in 72/105 (69%) and delineated the primary tumour in 38/105 (36%). This equated to an absolute EDR of 2.4% and a relative EDR of 2.9%. Missed lesions were concentrated in central and upper zones, whereas AI detections were more frequent in peripheral locations.

**Conclusions:** AI identified over one-third of retrospectively visible lung cancers that were missed at initial CXR reporting. Implementation as decision support could provide a modest but potentially meaningful increase in lung cancer detection in primary care.

**Advances in knowledge:** **I**n this large real-world UK study, AI modestly improved lung cancer detection on CXR, with complementary detection patterns to human readers, but performance remained limited in anatomically complex regions.

## Introduction

Lung cancer is the leading cause of cancer-related mortality worldwide, responsible for 1.8 million deaths annually (1). While low-dose computed tomography (CT) screening of high-risk populations reduces lung cancer mortality (2), chest radiography (CXR) remains the first-line investigation in primary care for patients with symptoms that might be caused by lung cancer. CXRs are inexpensive, readily available and have a sensitivity of 75-80% for the detection of lung cancer (3–4). This sensitivity reflects both the inherent limitations of the modality and human error in reporting and represents an opportunity for any interventions that could increase the detection on lung cancer on CXRs.

The UK Healthcare Safety Investigation Branch (HSIB) highlighted that missed opportunities to diagnose lung cancer on CXR remains a major patient safety concern, recommending national guidance for independent benchmarking of artificial intelligence (AI) tools to support CXR interpretation (5). Deep-learning models, particularly convolutional neural networks, have shown promise in detecting subtle radiographic abnormalities that may be overlooked by human readers (6). AI has been proposed as a second reader or triage tool to expedite lung cancer diagnosis, reduce reporting delays, and support workflow efficiency (7,8).

The potential clinical utility of AI in this setting is often expressed as an enhanced detection rate (EDR) — the proportion of additional cancers detected with AI support beyond standard reporting. Laboratory studies and systematic reviews have reported improvements in sensitivity when radiologists use AI (9–13), though the magnitude of benefit varies depending on algorithm performance, radiologist experience, and case mix. Importantly, these studies suggest AI and human readers may be complementary, detecting different subsets of cancers.

Despite encouraging early evidence, real-world evaluations remain limited. False positives could add unnecessary workload, while performance often varies with dataset characteristics and integration into local workflows (14–18). In practice, the margin for improvement may be smaller than laboratory studies suggest: a recent two-centre study of 25,000 CXRs found clinically significant misses by radiologists were rare, limiting the scope for AI benefit (19). Moreover, a NICE early value assessment concluded that current evidence was insufficient to support routine adoption of AI for lung cancer detection on CXR, calling for real-world studies addressing diagnostic accuracy, referral impact, and patient outcomes (20).

In response to these evidence gaps, we conducted a multicentre retrospective evaluation across nine NHS Trusts in our region. Using the Royal College of Radiologists’ national audit framework for missed lung cancers (21), we identified cases where a lung cancer was missed on CXR but was deemed to be retrospectively visible. We then assessed whether a commercially available AI tool could have identified these cases, thereby increasing detection. Our primary outcome was the enhanced detection rate (EDR), with secondary analyses examining AI sensitivity, error types, and anatomical distribution of missed lesions.

## Methods

### Study design and setting

We performed a retrospective, multicentre service evaluation across nine NHS Trusts across the region. The study followed the Royal College of Radiologists (RCR) audit template for “Missed Lung Cancers on Chest Radiographs” (21) and applied a five-step framework for clinical AI evaluation recommended in recent multi-society guidelines (22) (Supplementary Table 1).

### Case identification and inclusion criteria

Adult patients who underwent primary care–referred chest radiography between July 2019 and June 2022 were identified from institutional databases (∼280,000 CXRs). Cases were included if a new lung cancer diagnosis (ICD-10 C34) was confirmed within six months of the CXR. This yielded 1,600 CXRs linked to a confirmed lung cancer diagnosis.

### Identifying lung cancers missed on CXR

All 1,600 CXR reports were reviewed to classify each case as either *detected* (lesion described as suspicious for cancer or follow-up investigation recommended) or *missed* (lesion not described, or only non-specific findings reported, with no follow-up suggested). Reports were initially reviewed by a junior trainee and an experienced year-3 radiology registrar, with discrepancies resolved by a consultant radiologist with over 20 years’ experience.

Missed cases were further categorised as either *visible in retrospect* (lesion apparent once the tumour location was known from subsequent CT/PET imaging) or *not visible on CXR* (no abnormality apparent even in hindsight). This process identified 105 CXRs in which an abnormality corresponding to the cancer was retrospectively visible. Cases without a retrospectively visible abnormality were excluded from further AI analysis, to avoid overestimating performance.

### AI system

We used Annalise CXR (version 2.1; Annalise.ai, Australia), deployed via the Sectra Amplifier Platform. This commercially available deep-learning tool detects 124 thoracic abnormalities. AI analysis was performed on all 1,600 cancer-positive CXRs to estimate overall sensitivity and assess potential false negatives, and separately on the 105 missed-but-retrospectively-visible CXRs for enhanced detection rate (EDR) calculations. For the overall cohort, we recorded whether AI identified any actionable findings concerning for cancer (nodule, opacity, collapse). For the missed-but-retrospectively-visible cohort, we also recorded whether AI correctly delineated the primary tumour, based on CT/PET images.

### Expert review of missed cases

An independent consultant thoracic radiologist with over 20 years’ experience provided blinded review of the 105 missed cases and AI analysis. The reviewer initially interpreted each CXR without access to the original report or CT/PET findings, providing a management recommendation using the CX0–CX3 coding system (Supplementary Table 2). With access to the report and CT/PET findings, the reviewer then provided final confirmation of whether AI correctly detected the cancer (defined as delineating the abnormality corresponding to the primary tumour), the error type (observational vs interpretative), and lesion conspicuity (Likert scale, 1 = very subtle, 5 = very obvious).

### Outcomes

The primary outcome was the enhanced detection rate (EDR), defined as the proportion of confirmed cancers that were visible on CXR at baseline but not reported, which were then accurately delineated and identified by AI. We report both absolute EDR (AI-detected missed cancers ÷ total confirmed cancers) and relative EDR ([detection rate with AI – detection rate without AI] ÷ detection rate without AI × 100%).

Secondary outcomes included AI sensitivity across the full cancer cohort (n = 1,600), actionability of AI-flagged abnormalities (CX1–CX3), error categorisation and lesion conspicuity, and spatial distribution of detected versus missed lesions using a 3×3 anatomical grid overlay. Illustrative “WOW” cases were collected as case studies.

### Statistical analysis

Descriptive statistics were calculated for demographics and detection rates. The chi-squared test was used to compare lesion distribution across anatomical zones. Analyses were performed in R version 4.5.0.

### Ethical approval

This work was undertaken as a registered service evaluation within participating NHS trusts and did not require research ethics committee approval. All data were anonymised before analysis, in line with NHS information governance standards, GDPR, and the UK Policy Framework for Health and Social Care Research.

## Results

Of 1,600 CXRs linked to a lung cancer diagnosis, 1,296 were excluded because the initial report contained an actionable abnormality. Of the remaining 304, a tumour was retrospectively visible in 105 cases (6.6% of the total cohort; Figure 1). These CXRs formed the dataset for evaluating AI performance. Multiple CXRs from the same patient were included where available, with analyses performed at the image level.

**Figure 1.**
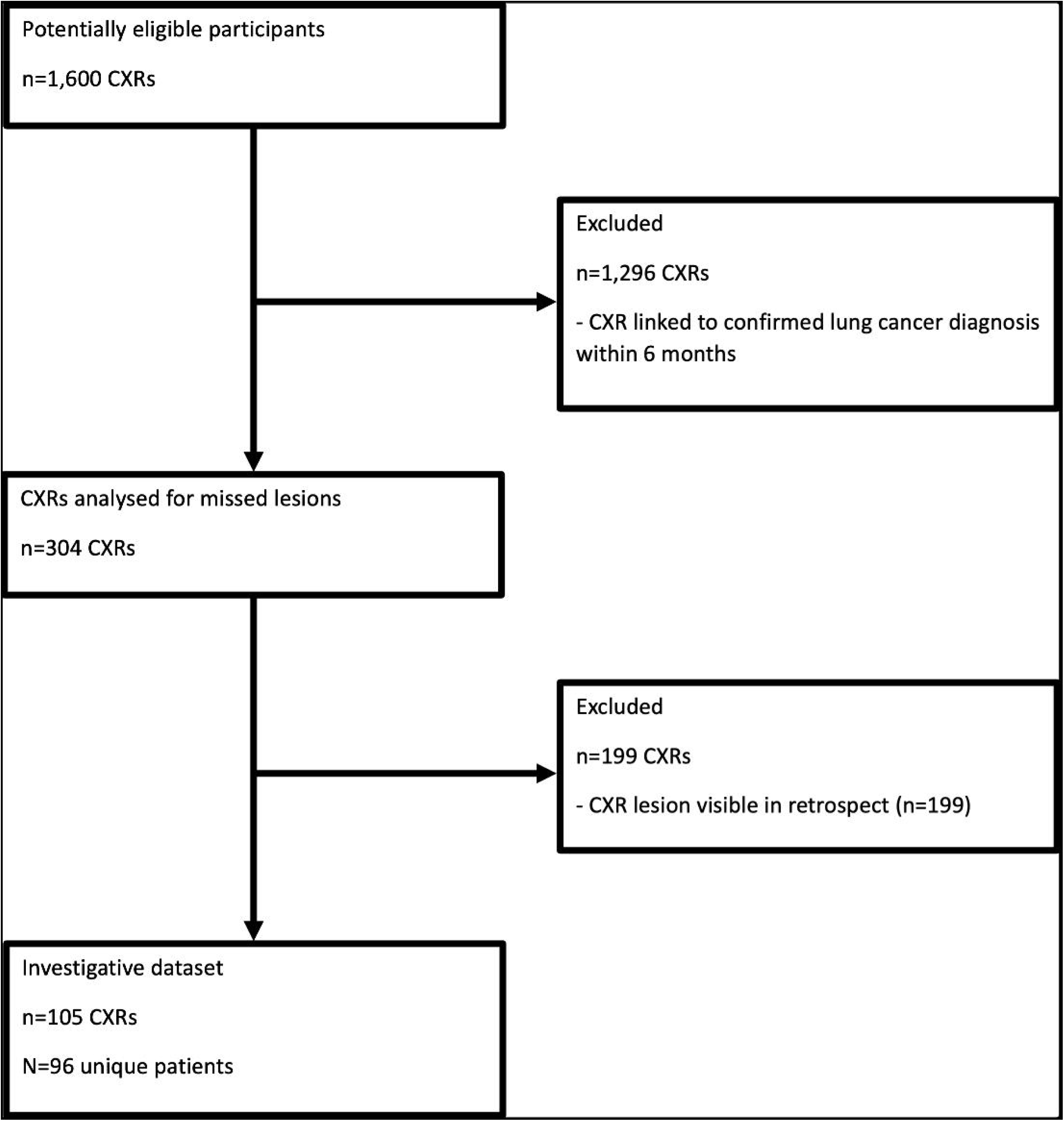
STARD diagram of participant selection. Among 1,600 GP-referred CXRs with lung cancer diagnosis, 1,296 had been positive on initial report and were excluded; 304 had no initial findings, of which 199 had no visible lesion on review, leaving 105 CXRs with a missed visible cancer for analysis.

### Study population

The final dataset comprised 105 CXRs from 96 patients with confirmed lung cancer that had been missed at initial reporting but were retrospectively visible. There was an equal gender distribution (49 males, 47 females). The mean age was 71 years (SD 9.8; range 47–88).

### Primary outcome

Within the 105 retrospectively visible cases, the AI flagged actionable abnormalities in 72/105 (69%). Following independent review, 38/105 (36%) were confirmed as true detections, where the AI delineated an abnormality at the site of the primary tumour. This equates to an absolute enhanced detection rate (EDR) of 2.4% (38/1600) and a relative EDR of 2.9% (83.9% vs 81.0%). If all AI-flagged abnormalities had led to further investigation, the theoretical absolute EDR would increase to 4.5% (72/1600).

### AI sensitivity across the full lung cancer cohort

Across all 1,600 cancer-positive CXRs, the AI flagged potentially relevant abnormalities (nodule, opacity, collapse) in 1,332 cases, corresponding to a sensitivity of 83%.

### Actionability of missed cases

Expert categorisation found that 82/105 (78%) contained actionable findings (CX1–CX3), while 44/105 (42%) were classified as highly suspicious for cancer (CX3).

### Illustrative ‘WOW’ cases

Examples of missed cancers detected by AI are shown in Figure 2. In these cases, the AI highlighted focal opacities or nodules corresponding to the primary tumour subsequently confirmed on CT/PET. The examples include an upper lobe opacity, a lower lobe mass, a solitary pulmonary nodule, and hilar lymphadenopathy.

**Figure 2.**
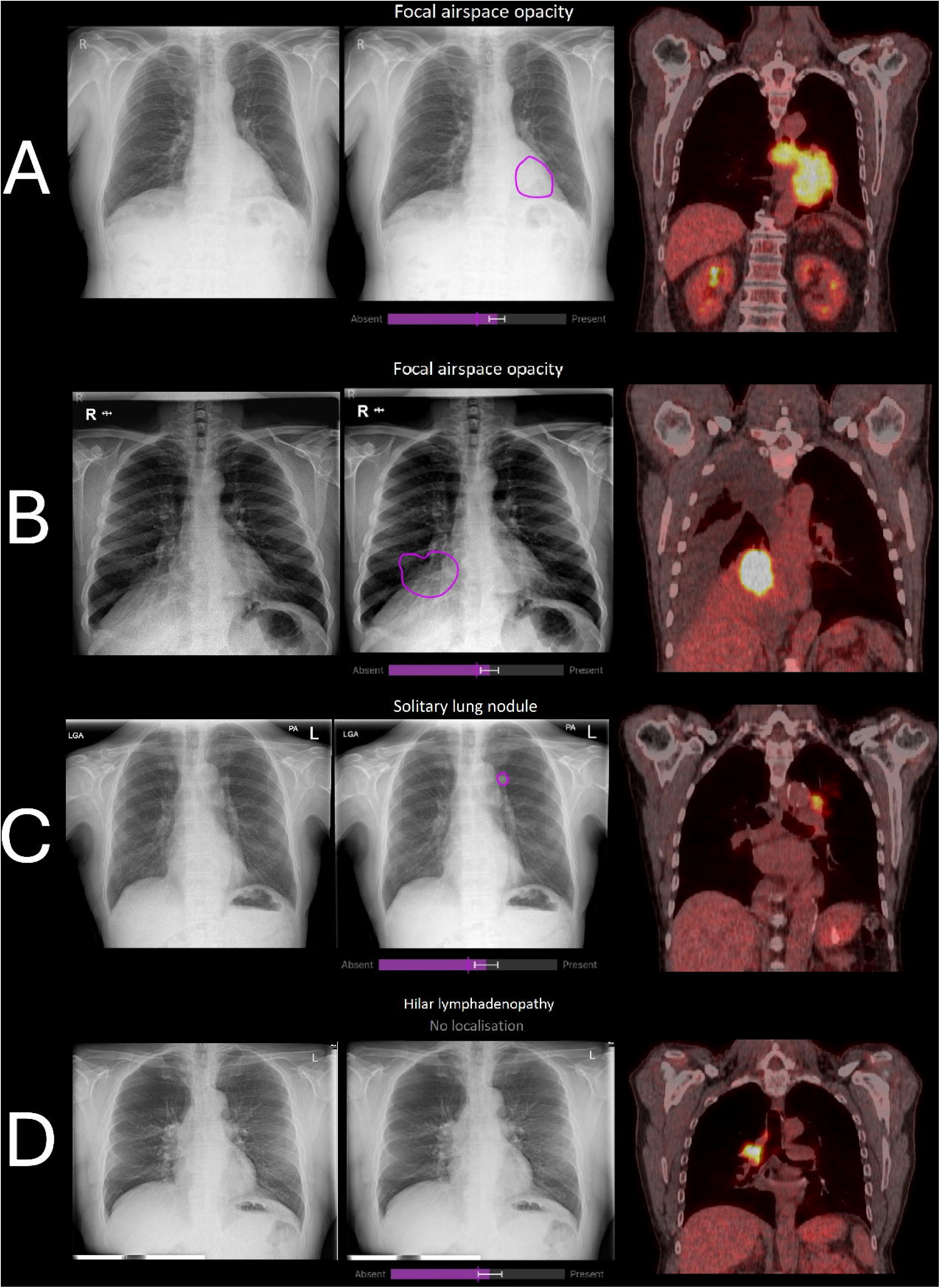
Examples of missed lung cancers detected by AI. Each row is a different patient: original CXR (left), AI annotated CXR (middle), and follow-up PET-CT (right) confirming the cancer. In these cases, the AI’s prompts (“focal opacity,” “solitary lung nodule,” etc.) corresponded to the abnormality and location where appropriate.

### Error types and conspicuity

Most missed cancers were observational errors (85/105, 81%), where an abnormality was present but not recognised. Interpretative errors were less common (20/105, 19%). AI detected 9/20 (45%) of interpretative errors and 29/85 (34%) of observational errors (Table 1). Conspicuity scores ranged from 1 (very subtle) to 5 (very obvious). Twenty-three cases (22%) were scored 1, 23 (22%) were scored 2, 14 (13%) were scored 3, 21 (20%) were scored 4, and 15 (14%) were scored 5. AI detection rates increased with conspicuity: 2/32 (6%) for score 1, 7/23 (30%) for score 2, 8/14 (57%) for score 3, 12/21 (57%) for score 4, and 9/15 (60%) for score 5 (Table 1).

**Table 1.** Annalise.AI detection rates in missed lung cancer cases (n = 105), stratified by error type and difficulty score. Difficulty scores (1–5) reflect expert ratings of conspicuity, where 1 = very subtle and 5 = very obvious. Error type was classified as interpretative (lesion seen but misinterpreted) or observational (lesion present but not recognised). Values are shown as “detected/total (%)”, where “detected” indicates cancers correctly flagged by AI.

| Difficulty | Interpretative<br>detected/total | % | Observational<br>detected/total | % | All cases<br>detected/total | % |
| --- | --- | --- | --- | --- | --- | --- |
| 1 (very subtle) | 2/7 | 29% | 0/25 | 0% | 2/32 | 6.3% |
| 2 | 3/6 | 50% | 4/17 | 24% | 7/23 | 30% |
| 3 | 1/3 | 33% | 7/11 | 64% | 8/14 | 57% |
| 4 | 2/3 | 67% | 10/18 | 56% | 12/21 | 57% |
| 5 (very obvious) | 1/1 | 100% | 8/14 | 57% | 9/15 | 60% |
| **Summary** | **9/20** | **45%** | **29/85** | **34%** | **38/105** | **36%** |

### Anatomical distribution of detection

Lesion locations were mapped to a 3×3 anatomical grid (Table 2 and Supplementary Figure 1). AI-detected cancers were most frequent in the middle-right (9/38, 24%), lower-right (9/38, 24%), and lower-left (5/38, 13%) zones. AI-missed cancers were concentrated in the middle-centre (20/67, 30%), middle-left (10/67, 15%), and lower-centre (10/67, 15%) zones. A chi-squared test suggested a trend towards different distributions (χ² = 14.70, df = 8, p = 0.065), although this was not statistically significant. Figure 3 illustrates these patterns, with heatmaps showing that human and AI misses were most frequent in anatomically complex regions such as the apices, hila, and retrocardiac zones.

**Figure 3.**
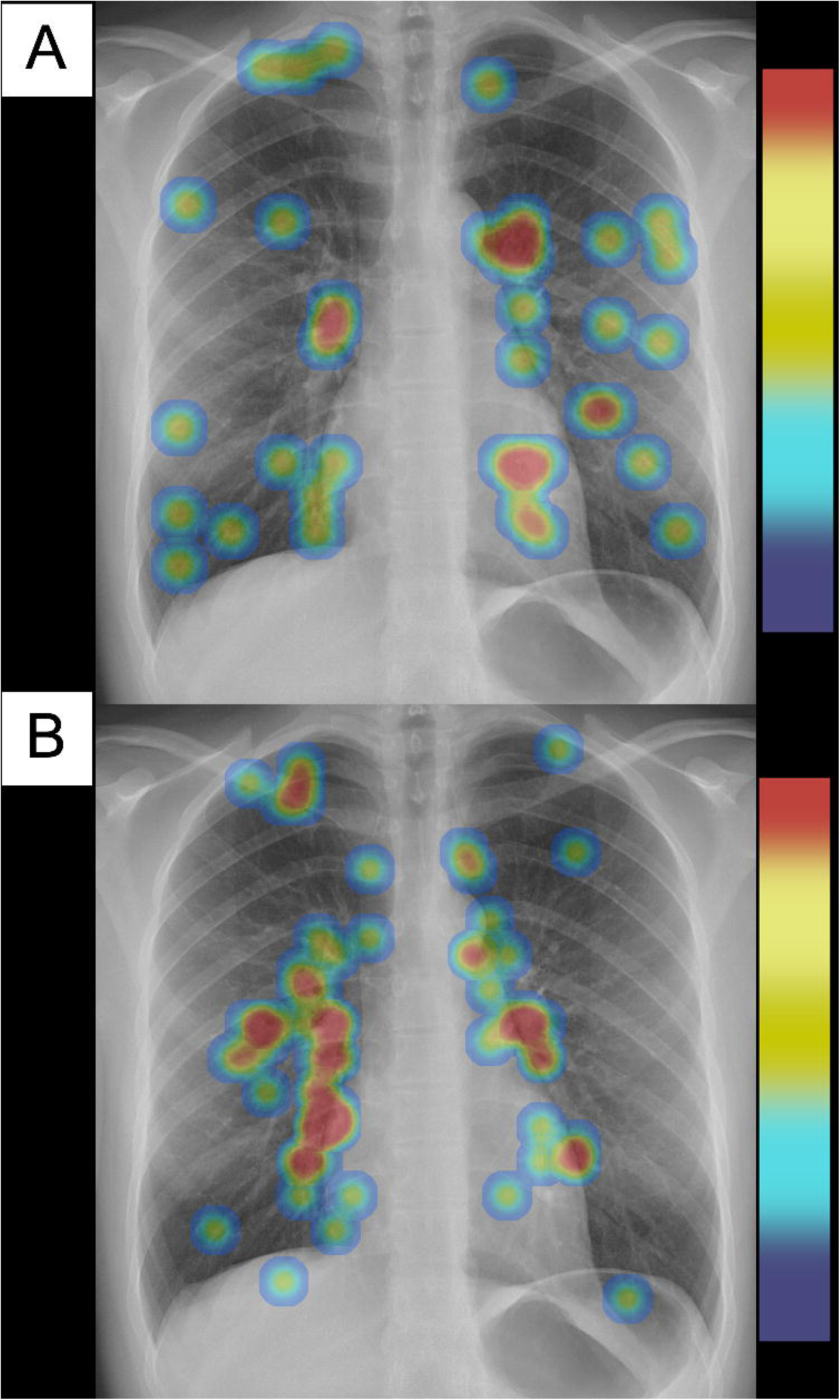
Heatmap of the anatomical locations of A) human missed and AI detected lung cancers on chest X-ray vs B) combined cases missed by humans and AI. Warmer colours (red/orange) indicate regions where cancers were more often missed. The apices, hila, perihilar mediastinum, and retrocardiac regions show the highest concentrations of misses.

**Table 2.** Distribution of cancers by anatomical zone for AI-detected and AI-missed cases. A 3×3 anatomical grid was applied to each chest radiograph, dividing the image into upper (U), middle (M), and lower (L) zones across left (L), centre (C), and right (R) regions. Counts represent the number of cancers located within each zone for AI-detected and AI-missed groups.

| Anatomical Zone | AI Detected | AI Missed | Total |
| --- | --- | --- | --- |
| UL | 2 | 5 | 7 |
| UC | 2 | 3 | 5 |
| UR | 0 | 2 | 2 |
| ML | 2 | 10 | 12 |
| MC | 5 | 20 | 25 |
| MR | 9 | 6 | 15 |
| LL | 5 | 5 | 10 |
| LC | 4 | 10 | 14 |
| LR | 9 | 6 | 15 |
| **Total** | **38** | **67** | **105** |

## Discussion

### Summary of key findings

In this real-world, multicentre evaluation, an AI detection system identified 36% of lung cancers that were retrospectively visible but initially missed on CXR. This translated into an absolute enhanced detection rate (EDR) of 2.4% and a relative EDR of 2.9% across 1,600 confirmed cases. Although modest in percentage terms, such gains may be meaningful when applied across large primary care populations.

### Comparison with existing evidence

Our findings align with previous meta-analyses showing that adding AI to human interpretation improves CXR sensitivity by 2–4 percentage points (9). They are also consistent with Topff et al. (19), who reported that clinically significant misses on CXR are uncommon, limiting the margin for AI to improve detection. The pattern of complementary performance is notable: AI detected cancers missed by humans, while humans detected cancers missed by AI, supporting the concept of combined reading.

Spatial analysis showed that AI was more effective in peripheral and lower zones. Performance was poorer in anatomically complex central regions such as the hila and mediastinum. These findings mirror established “blind spots” for radiologists (4,5) and suggest opportunities for targeted algorithm optimisation. Detection rates increased with conspicuity, from 6% for very subtle lesions to around 60% for the most obvious. However, the clinical value of AI may be greatest in intermediate lesions, where human perceptual errors are more common. Our “WOW” cases further illustrate how AI can highlight subtle lesions that were overlooked in the original report but subsequently confirmed on CT/PET.

### Clinical implications

A 2–3% improvement in detection may appear small, but in a high-incidence disease such as lung cancer, even incremental gains could reduce diagnostic delay for hundreds of patients annually in the NHS. This may improve access to curative treatment and survival, though this requires confirmation in prospective outcome studies (23). Importantly, AI flagged abnormalities in 69% of missed cases, while only 36% represented true primary tumour detections. If all AI-flagged abnormalities had prompted further investigation, the absolute EDR could theoretically rise to 4.5%, though this likely overestimates benefit. This distinction underscores the potential for AI to act as a triage tool, prompting earlier review rather than providing definitive diagnosis. Effective integration into workflow will therefore be critical. Over-reliance risks automation bias if clinicians dismiss cases not highlighted by AI, while excessive flagging could increase workload. Structured training and careful interface design are essential to maximise benefit while minimising harm.

### Strengths and limitations

Strengths of this study include the large, multicentre dataset, use of the RCR national audit framework, and comprehensive review across four specialist thoracic radiology clinicians, including two senior consultants with over 20 years’ experience each. Cross-sectional imaging with CT and PET provided a robust assessment of whether a lesion was retrospectively visible and whether the AI truly identified the primary tumour. By restricting analysis to cancers visible in hindsight, we focused on cases where AI could realistically add value.

Limitations include the retrospective design and restriction to cancer-positive CXRs, meaning specificity and false-positive rates could not be assessed. While we can describe the potential EDR, which estimates the added value of AI in CXR workflows, this does not capture AI-detected abnormalities that could prompt CT when lung cancer is not present. The impact of false positives is therefore an important area for future research. Furthermore, we can only hypothesise that AI identification of the primary tumour would have prompted reporting radiologists, who did not recommend further evaluation at the time, to refer for cancer investigations. The EDR should therefore be interpreted cautiously. The cohort was drawn from one UK region and may not generalise to other populations. Lesion size and conspicuity were not systematically measured, and downstream patient outcomes were not assessed. Finally, while health economic analysis was beyond scope, the modest EDR suggests that cost-effectiveness evaluations must weigh detection gains against the workload from follow-up imaging (7,25).

### Future directions

Randomised trials comparing radiologist performance with and without AI would provide the strongest evidence, but the scale required to detect a 2–3% gain would be prohibitive. With a cancer prevalence of ∼0.8% in primary care–referred CXRs, such a trial would need millions of participants to achieve adequate power, making such trials impractical. Pragmatic prospective service evaluations embedded in clinical workflow may therefore represent a more feasible path forward. Ongoing refinement of algorithms to address anatomically complex regions, together with studies on cost-effectiveness and impact on time to CT referral, will be essential for wider adoption.

## Conclusion

Artificial intelligence has the potential to modestly enhance lung cancer detection on primary care–referred chest radiographs by identifying a subset of cancers that were missed at initial reporting. Although the estimated gain is small (EDR ∼2–3%), the potential population-level benefit is meaningful. AI should be viewed as a supportive second reader rather than a diagnostic replacement. Careful workflow integration, user training, and continued optimisation for anatomically complex regions will be essential to realise the clinical value of AI in primary care lung cancer pathways.

## Data Availability

All data produced in the present study are available upon reasonable request to the authors

## Acknowledgements

The authors acknowledge Harrison.ai (formerly Annalise.ai) for providing access to the chest-radiograph tool evaluated in this study.

## Ethical approval

This project was registered locally as a service evaluation and conducted under the governance procedures of the participating NHS trusts. As no intervention or change to patient care was introduced, and all data were anonymised prior to analysis, formal research ethics committee approval was not required. The work was carried out in accordance with NHS information governance standards, GDPR, and the UK Policy Framework for Health and Social Care Research.

