## Supplementary material for "Enhanced Detection Rate of AI for Lung Cancer Detection on GP-Referred Chest X-rays: A Real-World Retrospective Evaluation": Supplmentary Figure 1

**Supplementary materials**

**Figure 1**

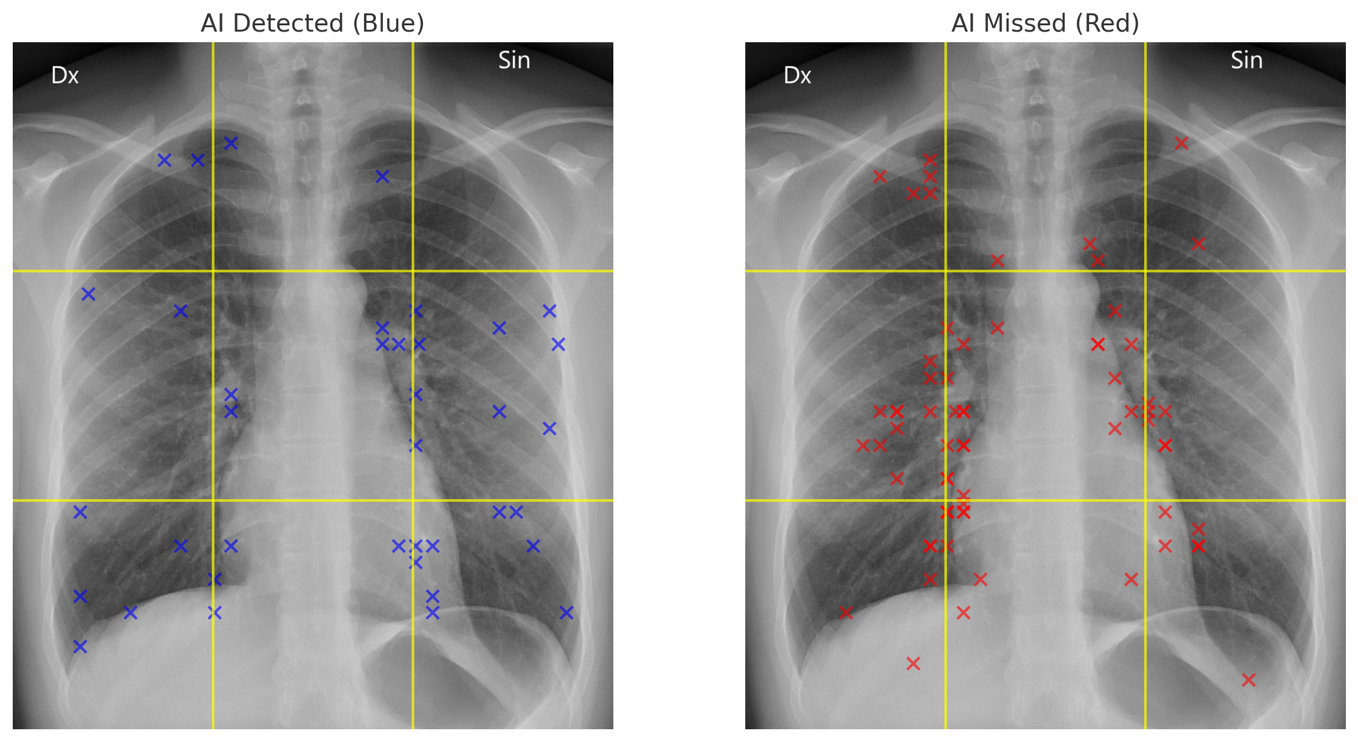
Figures showing 3×3 anatomical grid overlay on AI lesion heatmaps. Left image: Blue crosses are lesions detected by AI. Right image: Red crosses are lesions missed by AI. A 3×3 anatomical grid is overlaid to divide each image into upper (U), middle (M), and lower (L) zones across left (L), centre (C), and right (R) regions. This facilitates spatial comparison of lesion distribution patterns between detected and missed cases.
