## Supplementary material for "Enhanced Detection Rate of AI for Lung Cancer Detection on GP-Referred Chest X-rays: A Real-World Retrospective Evaluation": Supplmentary Table 1

**Supplementary materials**

**Table 1**

The Radiology Partners’ (RP) validation process emphasises continuous monitoring and real-world performance assessment, ensuring that AI tools remain effective and relevant over time. By focusing on metrics that matter to radiologists, RP aims to foster trust and encourage the adoption of AI technologies that genuinely enhance diagnostic accuracy and efficiency.

The five-step evaluation process included:

| Step | Description |
| --- | --- |
| 1. Review Model Accuracy | AI performance was retrospectively assessed on GP-referred CXRs to quantify its ability to flag actionable findings. |
| 2. Calculate Enhanced Detection Rate (EDR) | AI’s ability to detect previously missed lung cancers was evaluated against the original CXR reports. |
| 3. Identify 'WOW' Cases | AI-flagged cases were reviewed to determine high-impact instances where AI could have significantly altered management. |
| 4. Categorise Model Pitfalls | False positives and false negatives were analysed to assess AI’s limitations and areas for refinement. |
| 5. Summarise & Decide | The overall benefit and risk of AI integration were assessed, including training needs and automation bias mitigation. |
