## Supplementary material for "Enhanced Detection Rate of AI for Lung Cancer Detection on GP-Referred Chest X-rays: A Real-World Retrospective Evaluation": Supplmentary Table 2

**Supplementary materials**

**Table 2**

The following CX code definitions were used by the expert radiologist to categorise the chest radiographs by likelihood of malignancy.

- CX0: Nil acute – no planned follow up required (safety net only if persistent symptoms)

- CX1: Probable infection/transient findings – repeat CXR 6 weeks to check for resolution.

- CX2: Possible cancer- lower risk. For low dose non-contrast CT within 2 weeks (CATCH protocol)

- CX3: Probable cancer - higher risk. For contrast CT within 72 hours (RAPID protocol)
